# Evaluation of systemic immune response in patients with inflammatory complications of large joint replacements

**DOI:** 10.1101/2022.03.09.22271599

**Authors:** I.A. Mamonova, I.V. Babushkina, V.Yu. Ulyanov, A.S. Bondarenko, S.P. Shpinyak

## Abstract

We studied the features of systemic immunity in patients with inflammatory complications of their knee replacements and used the factor analysis to find immune characteristics associated with the progress of the pathological condition. The components indicative of changes related to the development of cell and humoral immune response in patients were isolated. Our findings present prospects for the development of new diagnostic techniques ensuring higher precision in searching for adequate surgical methods.

---

The number of patients who suffer from knee joint pathologies and need total joint replacements in Russia has been mounting yearly [6]. The multiplication of invasive interventions for artificial joint implantations leads to an increase in revisions. The aseptic loosening of the components of the implanted joint is one of the most frequent complications. The chronic inflammation caused by the response of the body’s immune system to the implant wear particles is believed to be the main reason for the implant aseptic loosening [5] while the septic inflammation is often induced by the chronic infection in the implantation area [1]. However, some studies claim that the diagnosis of periprosthetic inflammation is hindered with microbe biofilm formed by the pathogens; this results in making incorrect diagnoses leading to the wrong treatment strategy [2, 4]. Therefore, the investigation of mechanisms of aseptic inflammation progressing aimed at identifying pathogenic indicators of significant diagnostic value is a challenging line of research.

This research had the evaluation of systemic immune response and isolation the most important pathogenic indicators featuring the mechanisms of the immune system in patients with aseptic loosening of components in their knee implant as its objective.

## RESEARCH METHODS

The research involved 20 patients with aseptic loosening of components in their joint implants (16 men and 4 women) who received medical treatment in the Scientific Research Institute of Traumatology, Orthopedics and Neurosurgery, Federal State Budgetary Educational Institution of Higher Education ‘V.I. Razumovsky Saratov State Medical University’, the Russian Federation Ministry of Healthcare in 2019-2020. The patients’ mean age was 66.2 (56.31; 71.73) years old. Their diagnosis of *aseptic loosening of the joint implant components* was confirmed with the patients’ complaints of pain in the operated areas on exertion and at rest, as well as findings of X-ray and general examinations, bacterial culture tests, and the outcomes of surgical interventions for removal of the implant components. The exclusion criteria for the experimental group patients were rheumatoid arthritis, intraarticular corticosteroid injections administered no less than 3 weeks before the research. The control group included 20 apparently healthy volunteers (13 men and 7 women) of 69.8 (53.1; 78.7) years old with no musculoskeletal pathologies, allergies, autoimmune, infectious inflammatory diseases, hepatitis B, C, or HIV.

The immunological blood tests were run before surgical interventions. We detected the absolute counts of T-, B- and NK-cells in peripheral blood using monoclonal antibody kits BD Multitest 6-Color TBNK Reagent (BD, USA) in BD FACS Canto II cytometer (BD, USA). The quantitive evaluation of immunoglobulins in blood serum was performed with the immunoturbidimetric method in automatic open biochemical analyzer Sapphire-400» (Hirose Electronic Sistem Co., Japan) and Immunoglobulin A, M, G (FS) panel (DiaSys Diagnostic Systems GmbH, Germany). Cytokine serum concentrations were found with ELISA and Alfa-FNO-IFA-BEST, Interlejkin-6-IFA-BEST, Interlejkin-4-IFA-BEST, Interlejkin-10-IFA-BEST (Vektor-Brest Ltd., Russia) panels in Epoch device (BioTechUSA, USA).

The findings were processed using nonparametric statistical methods in MicrosoftExcel 2016 and Statistica 10.0. The testing for normality of distribution for quantitative indicators involved the Shapiro-Wilk test, and the significance of differences was evaluated with the Mann-Whitney U test. The factor analysis with the method of principal components was done in SPSS (11-2018) software suite. The Kaiser-Meyer-Olkin Measure criteria (the values should range 0.5 to 1) and the Bartlett test (the model is analyzable at P<0.05) were employed to determine the analyzability of the findings for factor analysis and the significance of the designed factor model. We used the method of principal components to find the factors and the Kaiser’s normal varimax to determine the best factor composition.

## RESEARCH RESULTS

The findings of the research for the evaluation of system immunity in the patients with aseptic loosening of the components in their joint implants are presented in Tables 1 and 2.

**Table 1.** Cell immunity indicators in the patients with aseptic loosening of the components in their joint implants

| Cell immunity indicators, cells/mcl |  |  |  |  |
| --- | --- | --- | --- | --- |
| CD3+CD19- | CD3+CD4+ | CD3+CD8+ | CD3- CD16+CD56+ | CD3-CD19+ |
| Control patients (n=20) |  |  |  |  |
| 1780.99<br>(1433.77;<br>1808.92) | 964.18<br>(949.28;<br>1123.82) | 658.41<br>(393.00;<br>745.70) | 166.18<br>(149.25;<br>288.66) | 321.55<br>(274.20;<br>340.01) |
| Patients with aseptic loosening of the components in their joint implants (n=20) |  |  |  |  |
| 1396.30<br>(1341.50;<br>1444.17)<br>$p=0.0006$ | 767.80<br>(712.10;<br>843.60)<br>$p=0.0001$ | 468.00<br>(438.90;<br>529.30)<br>$p=0.0019$ | 290.30<br>(108.40;<br>485.60) | 148.60<br>(138.20;<br>157.40)<br>$p=0.00001$ |

n – number of examined patients in the group; p – significance of differences in indicators to the controls.

**Table 2.** Immunoglobulin contents and cytokine levels in the patients with aseptic loosening of the components in their joint implants

| Indicators |  |  |  |  |  |  |
| --- | --- | --- | --- | --- | --- | --- |
| IgA<br>(pg/ml) | IgM<br>(pg/ml) | IgG<br>(pg/ml) | TNFα<br>(pg/ml) | IL-4<br>(pg/ml) | IL-10<br>(pg/ml) | IL-6<br>(pg/ml) |
| Control patients (n=20) |  |  |  |  |  |  |
| 101.00<br>(93.00;<br>115.50) | 153.7<br>(131.00;<br>181.30) | 1667.30<br>(1496.80;<br>1800.30) | 4.71<br>(4.01;<br>5.93) | 0.19<br>(0.07;<br>0.26) | 15.38<br>(11.96;<br>20.51) | 6.98<br>(4.93;<br>8.53) |

| Patients with aseptic loosening of the components in their joint implants (n=20) |  |  |  |  |  |  |
| --- | --- | --- | --- | --- | --- | --- |
| 138.5<br>(114.50;<br>165.00)<br>p=0.0154 | 129.1<br>(113.50;<br>137.00)<br>p=0.0005 | 140.90<br>(1244.00;<br>1525.00)<br>p=0.0133 | 10.1<br>(7.8; 12.5)<br>p=0.0001 | 0.2<br>(0.1;<br>0.3) | 10.60<br>(9.09;<br>12.90)<br>p=0.0001 | 20.30<br>(17.30;<br>25.50)<br>p=0.0001 |

n – number of examined patients in the group; p – significance of differences in indicators to the controls.

It was found that the absolute count of T-lymphocytes (CD3+CD19-) in peripheral blood of patients with aseptic loosening of the components in their joint implants significantly decreased as compared to the group of apparently healthy volunteers (p=0.0006) due to T-helpers (CD3+CD4+) (P=0.0001) as well as cytotoxic T-lymphocytes (CD3+CD8+) (P=0.0019). The absolute count of B-cells (CD3-CD19+) in peripheral blood in the controls was also higher (p=0.00001). No significant difference in the absolute count of NK-cells (CD3-CD16+CD56+) was found between the groups; however the patients with aseptic loosening of the components in their joint implants featured the upward trend of this indicator.

The aseptic loosening of the components in joint implants was accompanied with the increase of serum immunoglobulin A (p=0.0154) as well as the decrease of class M (P=0.0005) and G (P=0.0133) immunoglobulins.

We also noticed the increase of cytokines IL-6 (P=0.0001) and TNFα (P=0.0001) in serum of patients with aseptic loosening of their implants as compared to apparently healthy volunteers. The decrease in anti-inflammatory cytokine IL-10 (P=0.0001) along with the increase in the number proinflammatory cytokines was also noticeable. No significant differences in serum concentrations of IL-4 between the patients with aseptic loosening of the components in their joint implants and apparently healthy volunteers were observed.

We performed the factor analysis of the immunoassay findings in the patients with aseptic loosening of the components in their joint implants. This statistical method enables isolating the main component that consists of the complex of pathogenic factors. These factors affect each other stronger than others providing the explanation of the observed relations between the variables and detection of hidden regularities that suggest the specifics of immune response development in the patients with aseptic loosening of the components in their joint implants.

We designed the rotated component matrix (Table 3). The analysis revealed two factors (KMO=0.510 and Bartlett’s criterion=0.047<0.050) explaining 77.600 percent of all deviates in the totality of indicators. The contrast between the factors made 14 percent suggesting the differences between isolated components.

**Table 3.** The rotated component matrix of immune indicators in the patients with aseptic loosening of the components in their joint implants

| Indicators | Component |  |
| --- | --- | --- |
|  | 1 | 2 |
| Tumour necrosis factor (TNFα) | 0.879 | - |
| Absolute count of NK-cells (CD3- | 0.762 | - |
| CD16+CD56+) |  |  |
| Absolute count of T-lymphocytes<br>(CD3+CD19-) | -0.731 | - |
| Immunoglobulin M | - | 0.750 |
| Interleukin-4 (IL 4) | - | 0.669 |
| Proper values | 1.980 | 1.255 |
| Percent of the explained variance | 39.593 | 25.109 |

The first of the most significant matrix component with the factor loading of 1.98 featured a strong positive correlation of serum TNFα concentration (+0.879) with the absolute count of NK-cells (+0.762) while the correlation with the absolute count of T-lymphocytes in peripheral blood was negative (−0.731). The values of the first component suggest the most profound changes associated with the formation of cell immune response. It is known that TNFα participates in the regulation of inflammatory processes within the body as it provides lymphocyte migration to the focus of inflammation. The direct correlation of TNFα with NK-cell concentration suggests the role of this class of lymphocytes in the progress of the inflammatory process in the joint implant area. It is known that NK-cells cause lysis of foreign cells as well as altered endogenous cells of the body if the molecules of the main class I histocompatibility complex are absent on the surface regardless of the antibody content or complement system. No significant differences were observed between the absolute count of NK-cells in peripheral blood of the patients with aseptic loosening of the components in their joint implants and apparently healthy volunteers. However, the patients with aseptic loosening of the components in their joint implants featured the trend for increasing this indicator. The invert correlation of serum TNFα and absolute count of T-lymphocytes suggests active migration of these cells to the joint implant area as well as chronization of the inflammatory process.

The second matrix component combined 25.109 percent of dispersion and featured a strong correlation between the values of immunoglobulins M concentration (+0.750) and interleukine-4 (0.669) content in blood serum. The values of the second component present the most profound changes in the humoral component of the immune system in the patients with aseptic loosening of the components in their joint implants. It is known that B-lymphocytes are the main cells of the immune system ensuring humoral immune response due to the synthesis of immunoglobulins of various types. This process is regulated by Th2-lymphocytes and mediated by lymphocytes-produced cytokines IL-4, IL-5, and IL-13. The patients with aseptic loosening of their joint implants featured the decrease of B-lymphocytes in their peripheral blood as well as serum immunoglobulins of M and G classes along with normal IL-4 content suggesting the deterioration of the humoral immune system. This was probably related to the progress of allergic response to an implanted device. It’s worth noticing that immunoglobulin A levels in the patients with aseptic loosening of their joint implants turned out to be higher than that in the controls. Some authors claim this in combination with other factors may suggest osseous resorption processes [3].

The data resulting from our research offer the challenge of developing new integral methods ensuring timely diagnosis and choosing the right method of surgical management for patients with loosening of their knee joint implants.

## Data Availability

All data produced in the present study are available upon reasonable request to the authors

